# Machine learning models for predicting prostate cancer and clinically significant prostate cancer at biopsy: An updated analysis of an expanded Japanese cohort

**DOI:** 10.64898/2026.08.09.26360024

**Authors:** Takumi Takeuchi, Akira Nomiya

**Affiliations:** Department of Urology, Japan Organization of Occupational Health and Safety, Kanto Rosai Hospital, 1-1 Kizukisumiyoshi-cho, Nakahara-ku, Kawasaki 211-8510, Japan

**Keywords:** prostate cancer, prostate biopsy, machine learning, artificial neural network, cross-validation, Gleason score

## Abstract

**Background:** Our institution reported in 2019 on a multilayer artificial neural network (ANN) that predicted prostate cancer at biopsy in 334 patients. That model was trained with TensorFlow 1.x and scored at three fixed step counts, and hyperparameter selection was not kept separate from test evaluation. Here we revisit an expanded cohort from the same hospital using current machine-learning practice.

**Methods:** All biopsy episodes available in the institutional database were pooled (n = 526; 524 after one non-binary outcome code and one record with missing digital rectal examination [DRE] data were dropped). The seven predictors of the original report were retained: age, prior biopsy history, PSA, prostate volume, DRE, and MRI diffusion-weighted imaging findings in the peripheral and transition zones. Twenty-seven patients contributed more than one biopsy episode, so folds were built with patient-ID-grouped, stratified k-fold cross-validation (StratifiedGroupKFold; scikit-learn 1.8.0) at 3 and 5 folds, repeated over 10 random partitions, so that no patient had episodes in a training fold and a test fold at the same time. Four classifiers were compared: L2-regularized logistic regression, gradient boosting, random forest, and a shallow (single hidden layer) multilayer perceptron. Two outcomes were modeled: detection of any prostate cancer, and detection of clinically significant prostate cancer (Gleason score ≥ 7).

**Results:** Any-cancer prevalence was 55.7% (292/524); Gleason score ≥ 7 prevalence was 39.7% (208/524). Under repeated 5-fold cross-validation, gradient boosting discriminated best, both for any prostate cancer (mean AUC 0.826, 95% CI 0.823–0.830) and for Gleason score ≥ 7 (mean AUC 0.855, 95% CI 0.852–0.859). Random forest and logistic regression followed closely (AUC 0.81–0.85). The shallow multilayer perceptron did worse, and less consistently so (any-cancer AUC 0.671; Gleason score ≥ 7 AUC 0.742), falling below the deeper five-hidden-layer ANN of the 2019 report as well. Results with 3-fold cross-validation were essentially unchanged.

**Conclusions:** In the expanded cohort, regularized logistic regression, gradient boosting and random forest each discriminated prostate cancer at biopsy at least as well as the multilayer ANN reported earlier, with much simpler models and with hyperparameter tuning kept apart from performance estimation. At this sample size the shallow network gained nothing over the simpler alternatives. This is a preprint; the study has not undergone external peer review.

## Introduction

Prostate biopsy is indicated when prostate cancer is suspected, usually because serum prostate-specific antigen (PSA) is elevated, the digital rectal examination (DRE) is abnormal, or both. Better prediction before biopsy, of cancer in general and of clinically significant (higher Gleason score) disease in particular, would spare some men an invasive procedure that turns out to be unnecessary.

In 2019 we described a multilayer artificial neural network (ANN) built in TensorFlow, with two to five hidden layers of five neurons each, that predicted biopsy-detected prostate cancer in 334 men who had multiparametric MRI before transrectal 12-core biopsy [1]. That report laid out many combinations of variable-selection method, hidden-layer count and training-step count in one table and then reported the combination with the highest test-set AUC. Model selection was therefore not separated from the final performance estimate. Performance was also assessed on a single internal train/test split, with no external or temporal validation.

Larger studies have since applied conventional machine-learning or neural-network models to similar prebiopsy variables. Chiu et al. compared XGBoost, LightGBM, CatBoost, support vector machines, logistic regression and random forest using PSA, DRE and transrectal ultrasound findings, splitting their cohort temporally into a 2003–2017 training set and a 2017–2019 prospective validation set [2]. Parekh et al. built the Mount Sinai Prebiopsy Risk Calculator in 1,902 men (2,363 biopsies) and validated an ANN-based version externally, with AUCs of 0.90 for any prostate cancer and 0.92 for clinically significant prostate cancer in an independent European cohort [3]. Chen et al. and Checcucci et al. likewise reported machine-learning models, among them fuzzy-logic and ensemble approaches, that modestly outperformed PSA-based comparators for prostate cancer detection [4,5]. Set against the original 2019 report from our own institution, these studies share larger samples, simpler and more regularized models, and in several instances temporal or geographic external validation.

With that in mind, we re-analyzed data from the same institution, now grown to 526 biopsy episodes. Cross-validation was done at the patient level, non-neural-network baselines were included, and both an “any cancer” and a clinically significant (Gleason score ≥ 7) outcome were modeled.

## Methods

### Study design and setting

This is a retrospective, single-institution reanalysis of a biopsy database kept at the Department of Urology, Japan Organization of Occupational Health and Safety, Kanto Rosai Hospital, Kawasaki, Japan. The clinical protocol and the inclusion criteria are unchanged from our original report [1]. Men underwent 3-Tesla multiparametric MRI, then ultrasonography-guided, MRI-cognitively-targeted transrectal 12-core prostate biopsy: bilateral sampling of the peripheral zone from apex to base, two additional far-posterior/lateral peripheral-zone cores, and one transition-zone core per lobe. Seven patients with PSA 32–725 ng/mL, a positive DRE and positive MRI findings had sextant rather than 12-core biopsy, as in the original cohort; they are retained here without exclusion, again following the original report. Neither the biopsy protocol nor the method of MRI interpretation changed over the extended enrollment period. MRI reads remained binary positive/negative calls on the peripheral- and transition-zone T2-weighted and diffusion-weighted sequences, without formal PI-RADS scoring. PI-RADS scores were not analyzed in the present study because they had not been recorded systematically for a large share of the earlier cases in the database.

The 2019 report [1] analyzed 334 patients. The extraction used for the present reanalysis, drawn from the same institutional database, held 433 biopsy episodes (408 unique patients) at an earlier data-freeze and later grew to 526 biopsy episodes (497 unique patients; 27 patients contributing 2 or more episodes) by December 31, 2021. All 526 episodes entered the present analysis. None were withheld as a separate test set.

This study was conducted in accordance with the Declaration of Helsinki and approved by the Ethical Committee of the Japan Organization of Occupational Health and Safety, Kanto Rosai Hospital (protocol code 201730, approved March 8, 2018).

### Predictors and outcomes

Seven predictors were used, matching the Lasso- and stepwise-selected variable sets reported previously [1]: age at biopsy, number of prior biopsies, serum PSA, prostate volume, DRE finding, and binary MRI diffusion-weighted imaging findings in the peripheral zone and in the transition zone. Continuous variables were standardized (z-score).

Two binary outcomes were modeled. The first was detection of any prostate cancer on biopsy. The second was detection of clinically significant prostate cancer, defined as a maximum Gleason score of 7 or higher (International Society of Urological Pathology Grade Group ≥ 2) on any biopsy core, with men without cancer and men with Gleason score 6 (Grade Group 1) forming the reference group.

### Model development and validation

Four classifiers were compared: L2-regularized logistic regression, gradient boosting, random forest, and a shallow multilayer perceptron with a single hidden layer, in contrast to the two-to-five-hidden-layer ANN architecture of the original report. All four used the seven predictors listed above. The neural network and logistic regression pipelines standardized features internally, and class weighting (“balanced”) was applied to logistic regression and random forest.

Twenty-seven patients in the pooled cohort contributed more than one biopsy episode. Standard k-fold cross-validation risks leakage when episodes from one patient fall on both sides of a split, so we used scikit-learn’s StratifiedGroupKFold, grouping by patient identifier and stratifying by outcome, with k = 3 and k = 5 folds. For each fold configuration, hyperparameters were chosen by grid search nested inside the training portion of each outer fold: regularization strength for logistic regression; tree depth, learning rate and number of estimators for the ensemble methods; hidden-layer size and L2 penalty for the neural network. Performance was read off the corresponding held-out fold only, and no test data informed hyperparameter selection at any stage. A single arbitrary fold partition can shift the reported estimate, so we also repeated 3-fold and 5-fold cross-validation over 10 independent random partitions (distinct random seeds), using a fixed, representative hyperparameter set for each model taken from the single nested-grid-search run described above, and we report the mean area under the receiver operating characteristic curve (AUC) with its 95% confidence interval across those 10 repeats. This repeated-partition estimate reflects the variability attributable to fold assignment for a fixed model and hyperparameter set. It is not the sampling uncertainty of the underlying population AUC.

The design departs from the original report [1] in three respects. Hyperparameter selection and performance evaluation are kept strictly apart at every stage, so that no single “best” result is picked by inspecting test-set performance across a table of candidate configurations. Grouping by patient identifier blocks leakage from repeat-biopsy patients. And non-neural-network baselines (logistic regression, gradient boosting, random forest) are evaluated alongside the neural network under identical cross-validation.

### Software

All analyses were performed in Python 3.12.3 using scikit-learn 1.8.0, pandas 3.0.2, NumPy 2.4.4, and SciPy 1.17.1 [6].

### Statistical analysis

Discrimination was summarized as the area under the receiver operating characteristic curve (AUC). Continuous variables are given as median (range), categorical variables as n (%). AUCs were not compared between models by formal hypothesis testing; the comparisons here are descriptive, which suits the exploratory, single-institution, preprint character of this reanalysis.

## Results

### Cohort characteristics

Table 1 summarizes the 524 biopsy episodes (497 unique patients) left after one record with a non-binary outcome code and one record with missing DRE data were excluded. Median age at biopsy was 72 years (range 42–97), median PSA 9.2 ng/mL (range 0.5–3398.9) and median prostate volume 27.7 mL (range 7.2–274.4). DRE was positive in 130 episodes (24.8%), peripheral-zone DWI in 290 episodes (55.3%) and transition-zone DWI in 261 episodes (49.8%). Any prostate cancer was detected in 292 episodes (55.7%). Among the cancer-positive episodes, the maximum Gleason score was 6 in 84, 7 in 74, 8 in 89, 9 in 38 and 10 in 7, so the prevalence of Gleason score ≥ 7 (clinically significant) disease in the full cohort was 39.7% (208/524).

**Table 1.** Characteristics of the analyzed cohort (n = 524 biopsy episodes).

| Characteristic | Value |
| --- | --- |
| Biopsy episodes (unique patients) | 524 (497) |
| Age, years, median (range) | 72 (42–97) |
| PSA, ng/mL, median (range) | 9.2 (0.5–3398.9) |
| Prostate volume, mL, median (range) | 27.7 (7.2–274.4) |
| Prior biopsy $\geq 1$, n (%) | 78 (14.9) |
| DRE positive, n (%) | 130 (24.8) |
| MRI peripheral-zone DWI positive, n (%) | 290 (55.3) |
| MRI transition-zone DWI positive, n (%) | 261 (49.8) |
| Any prostate cancer, n (%) | 292 (55.7) |
| Gleason score 6, n (%) | 84 (16.0) |
| Gleason score 7, n (%) | 74 (14.1) |
| Gleason score 8, n (%) | 89 (17.0) |
| Gleason score 9, n (%) | 38 (7.3) |
| Gleason score 10, n (%) | 7 (1.3) |
| Gleason score $\geq 7$ (clinically significant), n (%) | 208 (39.7) |

### Prediction of any prostate cancer

Table 2 gives the mean AUC for each model, with the 95% confidence interval across 10 repeated partitions, under 3-fold and 5-fold patient-grouped cross-validation. Gradient boosting discriminated best (5-fold: AUC 0.826, 95% CI 0.823–0.830), with random forest (0.824, 0.822–0.826) and logistic regression (0.807, 0.804–0.810) close behind. The shallow multilayer perceptron trailed clearly (0.671, 0.668–0.675). Results with 3-fold cross-validation were essentially unchanged (Table 2). A single, non-repeated 5-fold run showed much wider fold-to-fold spread, with random forest fold AUCs ranging from 0.77 to 0.90; averaging over repeated partitions damped this considerably.

**Table 2.** Discrimination (AUC) for detection of any prostate cancer, patient-ID–grouped stratified cross-validation, repeated over 10 random partitions (n = 524).

| Model | 3-fold AUC (95% CI) | 5-fold AUC (95% CI) |
| --- | --- | --- |
| Logistic regression | 0.808 (0.806–0.810) | 0.807 (0.804–0.810) |
| Gradient boosting | 0.821 (0.816–0.826) | 0.826 (0.823–0.830) |
| Random forest | 0.821 (0.818–0.823) | 0.824 (0.822–0.826) |
| Shallow MLP (1 hidden layer) | 0.681 (0.677–0.686) | 0.671 (0.668–0.675) |

### Prediction of clinically significant prostate cancer (Gleason score ≥ 7)

Discrimination for Gleason score ≥ 7 was higher than for any-cancer detection in all four models (Table 3). Gradient boosting was again best (5-fold: AUC 0.855, 95% CI 0.852–0.859), with random forest and logistic regression both near 0.847. The shallow multilayer perceptron improved relative to the any-cancer outcome (0.742, 95% CI 0.727–0.757), but it remained the weakest of the four models and had the widest confidence interval, which points to greater sensitivity to fold partitioning.

**Table 3.** Discrimination (AUC) for detection of clinically significant prostate cancer (Gleason score ≥ 7), patient-ID–grouped stratified cross-validation, repeated over 10 random partitions (n = 524).

| Model | 3-fold AUC (95% CI) | 5-fold AUC (95% CI) |
| --- | --- | --- |
| Logistic regression | 0.846 (0.844–0.849) | 0.847 (0.846–0.849) |
| Gradient boosting | 0.851 (0.849–0.853) | 0.855 (0.852–0.859) |
| Random forest | 0.847 (0.845–0.849) | 0.847 (0.845–0.850) |
| Shallow MLP (1 hidden layer) | 0.711 (0.684–0.737) | 0.742 (0.727–0.757) |

As a robustness check, each model was additionally trained on the 433 biopsy episodes available at the earlier data-freeze and evaluated on the 124 biopsy episodes contributed by patients not present in that earlier extraction, a de facto temporal external-validation subset. Any-cancer AUCs there ran from 0.84 to 0.89 across models, in line with the pooled cross-validation estimates in Table 2, though numerically somewhat higher. The small size of the subset (n = 124) and its higher cancer prevalence (60.5%) are the likely explanation, rather than genuinely superior generalization. This analysis is exploratory, sits outside the primary pooled-cohort design reported above, and is included only to support the plausibility of the primary results.

### Discussion

In an expanded version of the cohort first reported in 2019, four alternative classifiers (regularized logistic regression, gradient boosting, random forest and a shallow neural network) discriminated any prostate cancer with AUCs of roughly 0.67 to 0.83, and clinically significant (Gleason score ≥ 7) prostate cancer with AUCs of roughly 0.71 to 0.86. Gradient boosting and random forest were marginally better than logistic regression, and all three beat the shallow neural network. None of these simpler, single- or few-hyperparameter models needed the two-to-five-hidden-layer architecture of the original ANN. The AUCs obtained here, 0.81 to 0.86, are comparable to the 0.71–0.76 reported for the multilayer ANN in the original 334-patient cohort [1], and for the clinically significant outcome somewhat higher.

Tree-based ensemble methods and regularized linear models that match or exceed a multilayer neural network on structured clinical variables, in a cohort of a few hundred patients, are not an unusual finding in this literature. Chiu et al., working with PSA, DRE and TRUS findings in a temporally validated design, found ensemble machine-learning methods modestly better than PSA and PSA density alone, without a clear advantage for deep architectures [2]. Parekh et al. reached considerably higher external-validation AUCs, 0.90 for any cancer and 0.92 for clinically significant cancer, with an ANN-based risk calculator [3]; their development cohort, however, was about seven times the size of ours (2,363 biopsies). Sample size rather than network depth per se may therefore be what mainly determines neural-network performance in this setting, a possibility our original report also raised when discussing its own modest ANN–logistic-regression differences [1].

Two methodological points separate the present reanalysis from the original report. First, the earlier comparison across 15 variable-selection/layer-count/step-count combinations picked the best-performing configuration by inspecting test-set AUCs directly (Table 4 of [1]), which invites optimistic bias from implicit test-set-based model selection; here the hyperparameter search is nested strictly within each training fold. Second, 27 patients in the pooled cohort contributed more than one biopsy episode, and ordinary k-fold cross-validation could place a patient’s episodes on both sides of a split. A direct comparison in this dataset showed only a small numerical difference between grouped and ungrouped cross-validation, which fits the modest fraction of repeat-biopsy patients, but grouping by patient is still the methodologically correct choice and should be adopted routinely.

Several limitations apply. This is a retrospective, single-institution analysis without a prospectively defined, non-overlapping external validation cohort; the exploratory temporal-validation subset (patients absent from the earlier data-freeze) was small (n = 124) and does not substitute for a formal external validation study. MRI was interpreted as binary positive/negative reads rather than by the now-standard PI-RADS system, which limits comparability with contemporary MRI-based risk calculators. Hyperparameters for the repeated cross-validation analysis were fixed from a single nested-grid-search run rather than re-optimized within every repeat; this is a pragmatic simplification, and a preliminary comparison suggested it did not materially change the results. Finally, the data come from a single Japanese occupational-health hospital, as in the original report, so generalizability to other populations and MRI protocols is unknown.

## Conclusion

In this updated, methodologically stricter reanalysis of an expanded single-institution cohort, simple regularized models, gradient boosting and random forest above all, discriminated prostate cancer and clinically significant prostate cancer at least as well as the multilayer artificial neural network reported in 2019, and did so without a deep architecture. For datasets of this size, simpler and more interpretable models, combined with patient-level cross-validation and with hyperparameter tuning kept separate from performance evaluation, are a reasonable default for prebiopsy prostate cancer prediction. Prospective, external and ideally multi-institutional validation is needed before any such model is used to guide biopsy decisions.

## Declarations

### Institutional review board statement

The study was conducted in accordance with the Declaration of Helsinki and approved by the Ethical Committee of the Japan Organization of Occupational Health and Safety, Kanto Rosai Hospital (protocol code 201730, approved March 8, 2018).

### Data availability

The datasets used during the current study are not publicly available due to ethical restrictions and the privacy of the participants.

### Use of artificial intelligence

Claude (Anthropic; Claude Sonnet 5, Opus 5), an AI assistant, was used to reanalyze the dataset, write and run the statistical and machine-learning code, generate the tables reported here, carry out the literature search cited in the Introduction and Discussion, and draft this manuscript, all under the direction and review of the authors. No other AI tool was used. The authors reviewed and verified the content of this manuscript and take full responsibility for it, including the accuracy of the analyses and citations.

### Author contributions

T.T.: conception, data curation, analysis oversight, manuscript drafting and revision; A.N.: data curation, critical revision of the manuscript.

